# Clinical coding of long COVID in primary care 2020-2023 in a cohort of 19 million adults: an OpenSAFELY analysis

**DOI:** 10.1101/2023.12.04.23299364

**Authors:** Alasdair D Henderson, Ben FC Butler-Cole, John Tazare, Laurie A Tomlinson, Michael Marks, Mark Jit, Andrew Briggs, Liang-Yu Lin, Oliver Carlile, Chris Bates, John Parry, Sebastian CJ Bacon, Iain Dillingham, William A Dennison, Ruth E Costello, Yinghui Wei, Alex J Walker, William Hulme, Ben Goldacre, Amir Mehrkar, Brian MacKenna, The OpenSAFELY Collaborative, Emily Herrett, Rosalind M Eggo

**Affiliations:** London School of Hygiene and Tropical Medicine, Keppel Street, London WC1E 7HT, UK; Bennett Institute for Applied Data Science, Nuffield Department of Primary Care Health Sciences, University of Oxford, OX2 6GG, UK; TPP, TPP House, 129 Low Lane, Horsforth, Leeds, LS18 5PX, UK; Patient and Public Involvement Steering Committee, London, UK; Centre for Mathematical Sciences, School of Engineering, Computing and Mathematics, University of Plymouth, Plymouth, UK

**Keywords:** Long COVID, vaccination, descriptive cohort

## Abstract

**Background:** Long COVID is the patient-coined term for the persistent symptoms of COVID-19 illness for weeks, months or years following the acute infection. There is a large burden of long COVID globally from self-reported data, but the epidemiology, causes and treatments remain poorly understood. Primary care is used to help identify and treat patients with long COVID and therefore Electronic Health Records (EHRs) of past COVID-19 patients could be used to help fill these knowledge gaps. We aimed to describe those with long COVID in primary care records in England.

**Methods:** With the approval of NHS England we used routine clinical data from over 19 million adults in England linked to SARS-COV-2 test result, hospitalisation and vaccination data to describe trends in the recording of 16 clinical codes related to long COVID between November 2020 and January 2023. We calculated rates per 100,000 person-years and plotted how these changed over time. We compared crude and minimally adjusted rates of recorded long COVID in patient records between different key demographic and vaccination characteristics using negative binomial models.

**Findings:** We identified a total of 55,465 people recorded to have long COVID over the study period, with incidence of new long COVID records increasing steadily over 2021, and declining over 2022. The overall rate per 100,000 person-years was 177.5 cases in women (95% CI: 175.5-179) and 100.5 men (99.5-102). In terms of vaccination against COVID-19, the lowest rates were observed in those with 3+ vaccine doses (103.5 [95% CI: 101.5-105]). Finally, the majority of those with a long COVID record did not have a recorded positive SARS-COV-2 test 12 weeks before the long COVID record.

**Interpretation:** EHR recorded long COVID remains very low compared and incident records of long COVID declined over 2022. We found the lowest rates of recorded long COVID in people with 3 or more vaccine doses. We summarised several sources of possible bias for researchers using EHRs to study long COVID.

## Background

Some people experience prolonged symptoms for weeks or months following acute SARS-COV-2 infection. This sequelae is known as long COVID, which is probably best currently conceptualised not as a single disease entity but as a classification designed to include all individuals who develop persistent symptoms following acute SARS-CoV-2 infection. This classification likely represents multiple underlying syndromes including cardiovascular, thrombotic and cerebrovascular disease, myalgic encephalomyelitis/chronic fatigue syndrome and dysautonomia (1,2), each with distinct pathophysiologies and prognoses (3–7) and in some individuals these symptoms can be long-lasting and severe (8–11). The heterogeneity within the classification contributes to inconsistent definitions of long COVID across studies with resulting wide variation in estimated prevalence (12–16) and risk of developing long COVID following SARS-COV-2 infection (17–21).

Given this uncertainty, more research of the causes and consequences of long COVID is necessary (22). Electronic health records (EHRs) are a possible data source for this research and they have become critical in healthcare research (23–26), therefore careful analysis of EHRs could present an opportunity to better understand long COVID (27–31). However, common problems with EHR data include diagnostic accuracy, inconsistent coding, missing data and ascertainment bias (24,32,33). In the UK, diagnostic and referral codes for long COVID have been available for General Practitioners (GPs) in the UK since winter 2020, along with guidelines on use of these codes from the National Institute for Clinical Excellence (NICE guideline [NG188]).

We have previously summarised the early clinical coding of long COVID up to May 2021 and shown very low recording of long COVID (27). However, since then, different SARS-COV-2 variants have emerged and many COVID-19 vaccines administered, and there have likely been changes in coding practices. It is vital to understand any potential differences in coding before we can use EHRs to answer more complex research questions about long COVID. We therefore set out to comprehensively describe the incidence of GP-recorded long COVID, including the demographic and clinical characteristics of individuals with a long COVID code in England using OpenSAFELY, and associations with infection and vaccination history.

## Methods

### Data Source

We used a database of 19 million adults in England, whose primary care records are managed by the GP software provider TPP SystmOne. We accessed these data through the OpenSAFELY platform, where all data were linked, stored and analysed securely (https://opensafely.org/). Data, including coded diagnoses, medications and physiological parameters, are pseudonymised. No free text data are included. The following linked data were also used for this study: patient-level COVID-19 vaccination status via the National Immunisation Management System (NIMS); in-patient hospital spell records via NHS Digital’s Hospital Episode Statistics (HES); national coronavirus testing records via the Second Generation Surveillance System (SGSS); Detailed pseudonymised patient data are potentially re-identifiable and therefore not shared.

### Study population

We included all individuals aged 18-100 years and registered with a general practice that uses TPP SystmOne software on or after 1 November 2020, the date that long COVID SNOMED codes become available. SNOMED codes are a dictionary of computer-readable codes relating to clinical terms. Participants were followed up from the beginning of their registration plus 90 days to account for onboarding of EHR records after registering at a new practice. Participants were then followed until the earliest of: EHR record of long COVID; end of registration with the same general practice; death; or 31st January 2023 (**Figure <u>S1-S3</u>** (34)).

We also analysed vaccination coverage after a long COVID record, however the primary cohort ends follow-up at the time of recorded long COVID. We therefore developed a secondary cohort that includes only those with a record of long COVID and follows up until January 2023 or loss to follow-up, and we summarised vaccine coverage in this cohort as of January 2023 (**Supplementary Methods**).

### Outcomes

The primary outcome was the first GP record of long COVID defined by 15 SNOMED codes, as used previously (27,35–37) which are split between two groups of codes: diagnosis and referral (**Table 1**). Records were searched for a diagnosis code first, if no code existed then we searched for a referral code. If neither code existed then the individual was classified as not having long COVID.

**Table 1:** List of codes used to identify long COVID in the EHR record.

| SNOMED CT code | Description |
| --- | --- |
| *Diagnosis codes* |  |
| 1325161000000102 | Post-COVID-19 syndrome |
| 1325181000000106 | Ongoing symptomatic disease caused by severe acute respiratory syndrome coronavirus 2 |
| *Referral codes* |  |
| 1325021000000106 | Signposting to Your COVID Recovery |
| 1325031000000108 | Referral to post-COVID assessment clinic |
| 1325041000000104 | Referral to Your COVID Recovery rehabilitation platform |
| 1325051000000101 | Newcastle post-COVID syndrome Follow-up Screening Questionnaire |
| 1325061000000103 | Assessment using Newcastle post-COVID syndrome Follow-up Screening Questionnaire |
| 1325071000000105 | COVID-19 Yorkshire Rehabilitation Screening tool |
| 1325081000000107 | Assessment using COVID-19 Yorkshire Rehabilitation Screening tool |
| 1325091000000109 | Post-COVID-19 Functional Status Scale patient self-report |
| 1325101000000101 | Assessment using Post-COVID-19 Functional Status Scale patient self-report |
| 1325121000000105 | Post-COVID-19 Functional Status Scale patient self-report final scale grade |
| 1325131000000107 | Post-COVID-19 Functional Status Scale structured interview final scale grade |
| 1325141000000103 | Assessment using Post-COVID-19 Functional Status Scale structured interview |
| 1325151000000100 | Post-COVID-19 Functional Status Scale structured interview |

To ensure our cohort produced results consistent with previous literature, we included a control outcome for which we knew the expected direction of effect estimates. We used hospitalisation with COVID-19 (**Supplementary Methods)** which should have a negative association with SARS-CoV-2 vaccination based on previous research (38,39). To account for the gap between SARS-CoV-2 infection and the recording of long COVID, we only analysed COVID-19 test results and hospitalisations >12 weeks before the end of follow up. Data on SARS-CoV-2 test results were available from the SGSS.

### Stratifiers

COVID-19 vaccination status was the only time-updated covariate and was categorised in two ways: i) follow-up was divided by the number of vaccine doses received (0, 1, 2, 3+); ii) participants were categorised as having received an mRNA-based vaccine (Pfizer (Comirnarty), Moderna (Spikevax)) for their first immunisation, or a non-mRNA vaccine. Only vaccine doses greater than 14 weeks before the end of follow up were included to account for the gap between immunisation and protection (2 weeks) and development of long COVID symptoms (12 weeks).

All other covariates were defined at baseline, the start of a valid registration with a GP in the study period. Age (18-29, 30-39, 40-49, 50-59, 60-69, 70+) and sex. NHS region (9 regions in England), and index of multiple deprivation (IMD) quintiles were based on the address of each participant. Ethnicity (categorised as white, Black, South Asian, mixed, Other) (40), and those at high-risk of complications from COVID-19 (41) were assessed from primary care records. The presence of fifteen chronic comorbidities identified as increasing the risk of severe COVID-19 disease from previous research (15) were defined using primary care records at baseline and categorised (0, 1, 2+), the full list of comorbidities is available in the Supplementary Methods. Finally, we defined two binary “probable shielding” variables, one for those at “high-risk” of complications from SARS-CoV-2 infection, and one for those at low/moderate-risk. Both shielding variables were based on the presence of the corresponding SNOMED code (41) and are defined as “probable” as they are a proxy for shielding behaviour in those with the code recorded.

## Statistical methods

### Primary analysis

We estimated the crude rate of long COVID per 100,000 person years and 95% confidence intervals for each level of each stratifier listed (**Supplementary Methods**). All counts for presentation were rounded to the nearest 5 for and counts lower than 10 were redacted to ensure results are non-disclosive.

To compare rates between levels of each stratifier while partially adjusting for confounding, we developed negative binomial models for age category, sex and vaccination. All models were adjusted for age, sex, NHS region and dominant SARS-CoV-2 variant period (wildtype/alpha, 1 November 2020 - 16 May 2021; Delta, 16 May 2021 - 1 December 2021; Omicron, 1 December 2021 - 31 Jan 2023 (42)) to estimate rate ratios.

### Secondary analyses

We presented the incidence of long COVID recording by specific SNOMED codes and compared these to national recording of SARS-CoV-2 dynamics. We also described the reported SARS-CoV-2 history of people with EHR records of long COVID in a Sankey diagram, from SARS-CoV-2 test status, to COVID-19 hospitalisation to EHR recorded long COVID. We then compared the characteristics of those with and without a recorded SARS-CoV-2 positive test before their record of long COVID.

We expanded the negative binomial models further by running separate models for each of the three variant periods to analyse the consistency of these associations across the pandemic. We also conducted a sensitivity analysis of our vaccine definition by including vaccinations >14 weeks before end of follow-up (main analysis), to results from >16 weeks and >26 weeks.

Finally, we used the secondary cohort and calculated the percentage of people with either 0, 1, 2, or 3+ vaccine doses at the end of the study, stratified by whether they had a long COVID record previously and by age group.

### Software and Reproducibility

All data management and analysis code is available on GitHub (https://github.com/opensafely/openprompt-vaccine-long-covid). Data management and analysis was performed using the OpenSAFELY software libraries and Python 3. All analysis was conducted in R version 4.2.1 (43). All codelists used to define conditions and variables are openly available online at www.OpenCodelists.org for inspection and re-use. Definitions used in this study reuse codelists developed for published studies (27,38,44,45).

### Patient and Public Involvement

The OpenSAFELY platform team has developed a publicly available website https://opensafely.org/ which describes the platform in language suitable for a lay audience. This specific study design was developed with input from our patient and public advisory panel of three through regular meetings, and following feedback from two large workshops hosted in January and September 2023.

## Results

### Variation in incidence of long COVID recording in England

We analysed data from 19,462,260 adults in England between November 2020 and January 2023 with a median follow up time of 2.2 years. There was an even split of men and women, and 70% of the cohort were recorded as white ethnicity. Most of the cohort lived in the East Midlands (17%), East (23%), South West (14%) and Yorkshire & the Humber (14%) reflecting where SystmOne is used. Over a third of the cohort had at least one chronic comorbidity (**Table 2**).

**Table 2:** Baseline cohort characteristics. Figures shown are n (%) for binary and categorical variables, median (25% - 75% percentile) for continuous variables.

| Variable | Level | Cohort summary |
| --- | --- | --- |
| Total |  | 19,462,080 |
| Follow-up start (year) | 2020 | 17,824,820 (91.6%) |
|  | 2021 | 884,790 (4.5%) |
|  | 2022 | 675,120 (3.5%) |
|  | 2023 | 77,530 (0.4%) |
| Sex | male | 9,720,385 (49.9%) |
|  | female | 9,741,875 (50.1%) |
| Age (IQR) |  | 48 (33-63) |
| Age category | 18-29 | 4,078,305 (21%) |
|  | 30-39 | 3,450,885 (17.7%) |
|  | 40-49 | 3,129,670 (16.1%) |
|  | 50-59 | 3,271,525 (16.8%) |
|  | 60-69 | 2,527,420 (13%) |
|  | 70+ | 3,004,460 (15.4%) |
| Ethnicity | White | 13,586,060 (69.8%) |
|  | Mixed | 234,525 (1.2%) |
|  | South Asian | 1,356,505 (7%) |
|  | Black | 470,675 (2.4%) |
|  | Other | 514,690 (2.6%) |
|  | (Missing) | 3,299,805 (17%) |
| Region | London | 1,559,650 (8%) |
|  | East Midlands | 3,298,290 (16.9%) |
|  | East | 4,403,440 (22.6%) |
|  | North East | 904,835 (4.6%) |
|  | North West | 1,702,730 (8.7%) |
|  | South East | 1,288,965 (6.6%) |
|  | South West | 2,791,195 (14.3%) |
|  | West Midlands | 789,260 (4.1%) |
|  | Yorkshire and The Humber | 2,709,565 (13.9%) |
|  | (Missing) | 14,340 (0.1%) |
| IMD (quintile) | 1 (most deprived) | 3,508,485 (18%) |
|  | 2 | 3,677,465 (18.9%) |
|  | 3 | 3,990,465 (20.5%) |
|  | 4 | 3,761,210 (19.3%) |
|  | 5 (least deprived) | 3,499,345 (18%) |
|  | (Missing) | 1,025,285 (5.3%) |
| Comorbidities | 0 | 12,441,695 (63.9%) |
|  | 1 | 4,966,470 (25.5%) |
|  | 2+ | 2,054,095 (10.6%) |
| Probably shielding (high risk group) |  | 957,765 (4.9%) |
| Probably shielding (Low/moderate risk group) |  | 463,750 (2.4%) |

We identified 55,465 recorded codes for long COVID, 20,025 of which were diagnosis codes, with the remaining 35,440 referral codes (**Table <u>S1</u>**). The number of newly recorded (incident) long COVID cases increased steadily over 2021, before peaking in January 2022 and declining steadily for the following 12 months (**Figure 1**). This pattern was more pronounced when we included referral codes for long COVID in our outcome definition. The dynamics of which specific codes were used has changed over time. Initially, the two diagnosis codes contributed approximately half of all recorded codes. Since mid-2022 however, the majority of new codes have been referrals to post-COVID assessment clinics (**Figure 1B**). Clinical codes for long COVID became available at a similar time as the rollout of SARS-CoV-2 vaccination. Therefore, as the number of incident long COVID records increases over time, they are increasingly recorded in those that have been vaccinated (**Figure 1C**), which is consistent when stratified by sex (**Figure <u>S4</u>**).

**Figure 1:**
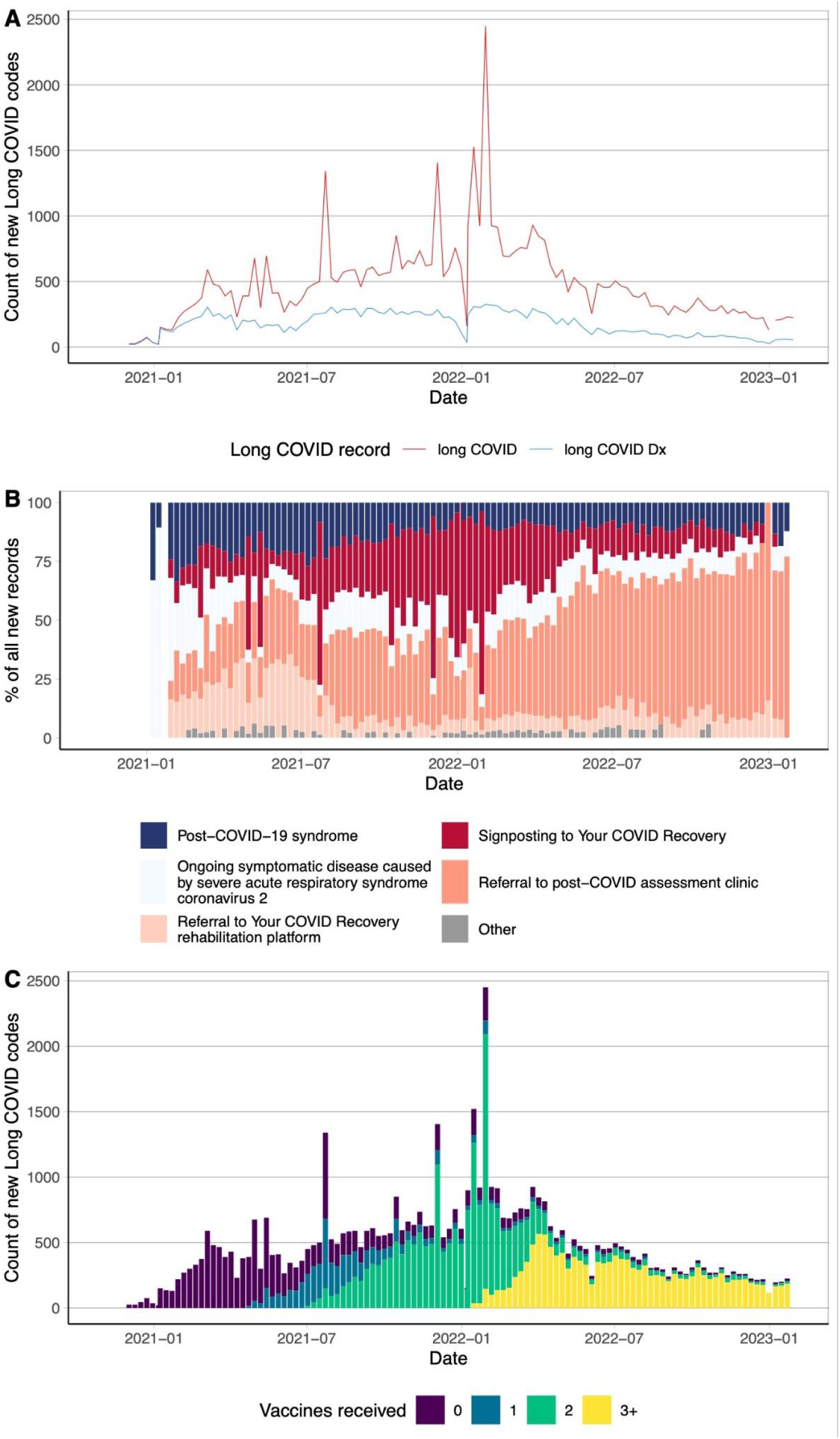
*Dynamics of long COVID recording in EHRs. A: weekly count of long COVID codes (any long COVID code, red; of which were diagnosis codes, blue). B: weekly proportion of the 5 most common long COVID codes amongst all new long COVID codes recorded that week. C: weekly count of all long COVID codes stratified by the number of vaccine doses received ≥14 weeks prior to the long COVID code*

The weekly pattern appeared to mask more marked variation in the recording of long COVID codes and we identified certain dates with large spikes (Figure 2). The main cause of these outliers in the time series appeared to be the use of one SNOMED code (“Signposting to Your COVID Recovery”) with three notable spikes in July 2021, December 2021 and January 2022. The pattern of long COVID recording over time did not visually appear to reflect the dynamics of positive SARS-COV-2 at a national level (**Figure <u>S5</u>**).

**Figure 2:**
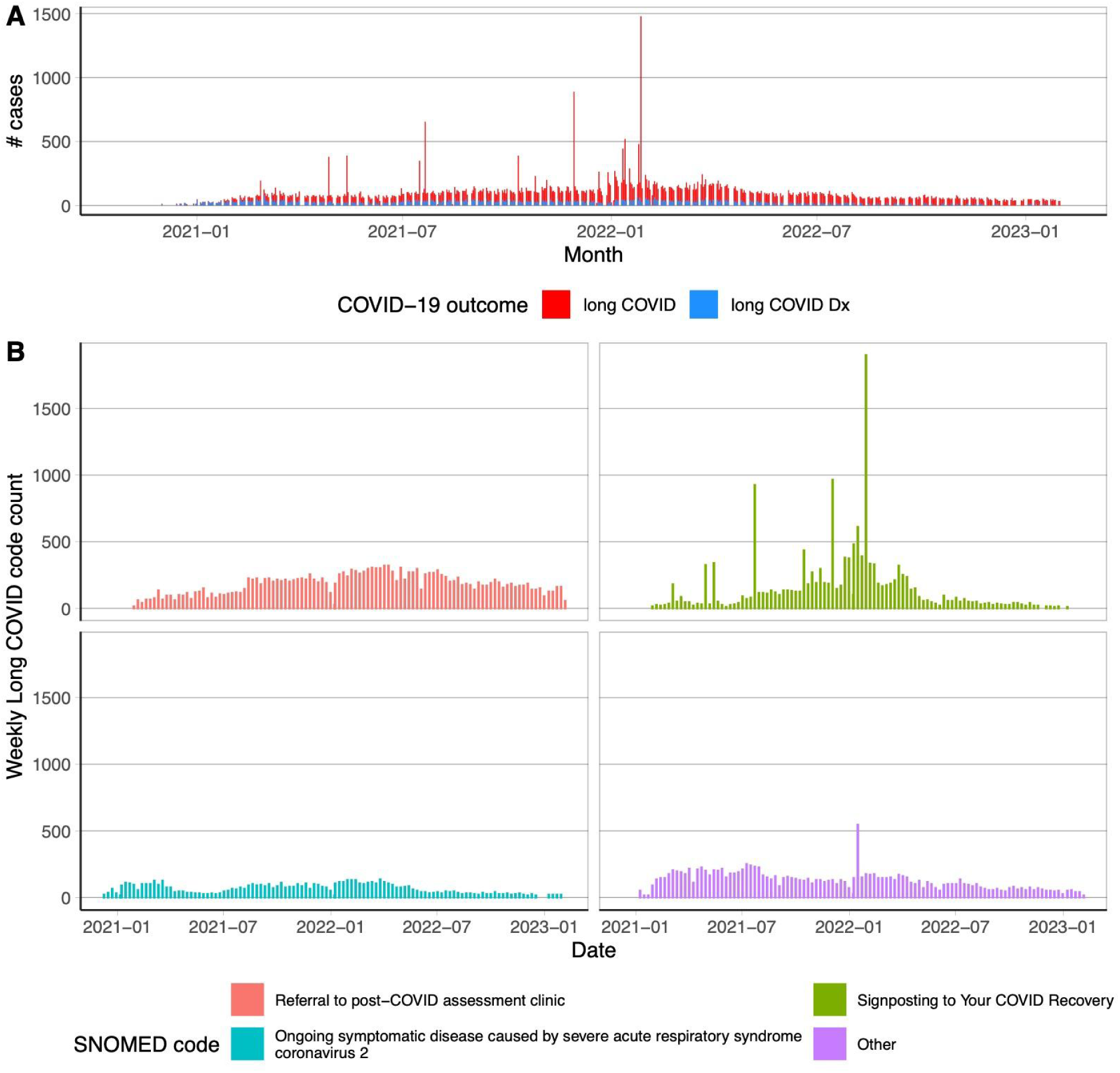
*primary care coding of long COVID codes over time. A: daily counts of any long COVID code (red) and long COVID diagnoses only (blue). B: weekly counts of the three most common long COVID codes in primary care, and the remaining codes grouped as “other”. Counts less than 10 are suppressed*.

### Recorded long COVID rates vary between population groups

Crude rates of long COVID coding were highest for women, ages 40-60, white ethnicity, those with at least one comorbidity and people who were shielding because they were at high-risk of complications from COVID-19 (Figure 3). The crude rate of long COVID records were lowest in those with 3+ vaccine doses, and were lower for those who received an mRNA-based vaccine as their first dose. However, the raw rate of long COVID codes was higher in those with one or two doses of the vaccine, reflecting the timing of vaccine rollout and long COVID code availability (Figure 1C). Finally, some patterns in the crude rates of EHR recorded long COVID were dependent on whether referral codes are included in the definition. Notably, we found that long COVID codes were more likely to occur in people living in less deprived areas, however this association did not hold when long COVID diagnosis codes only were analysed.

**Figure 3:**
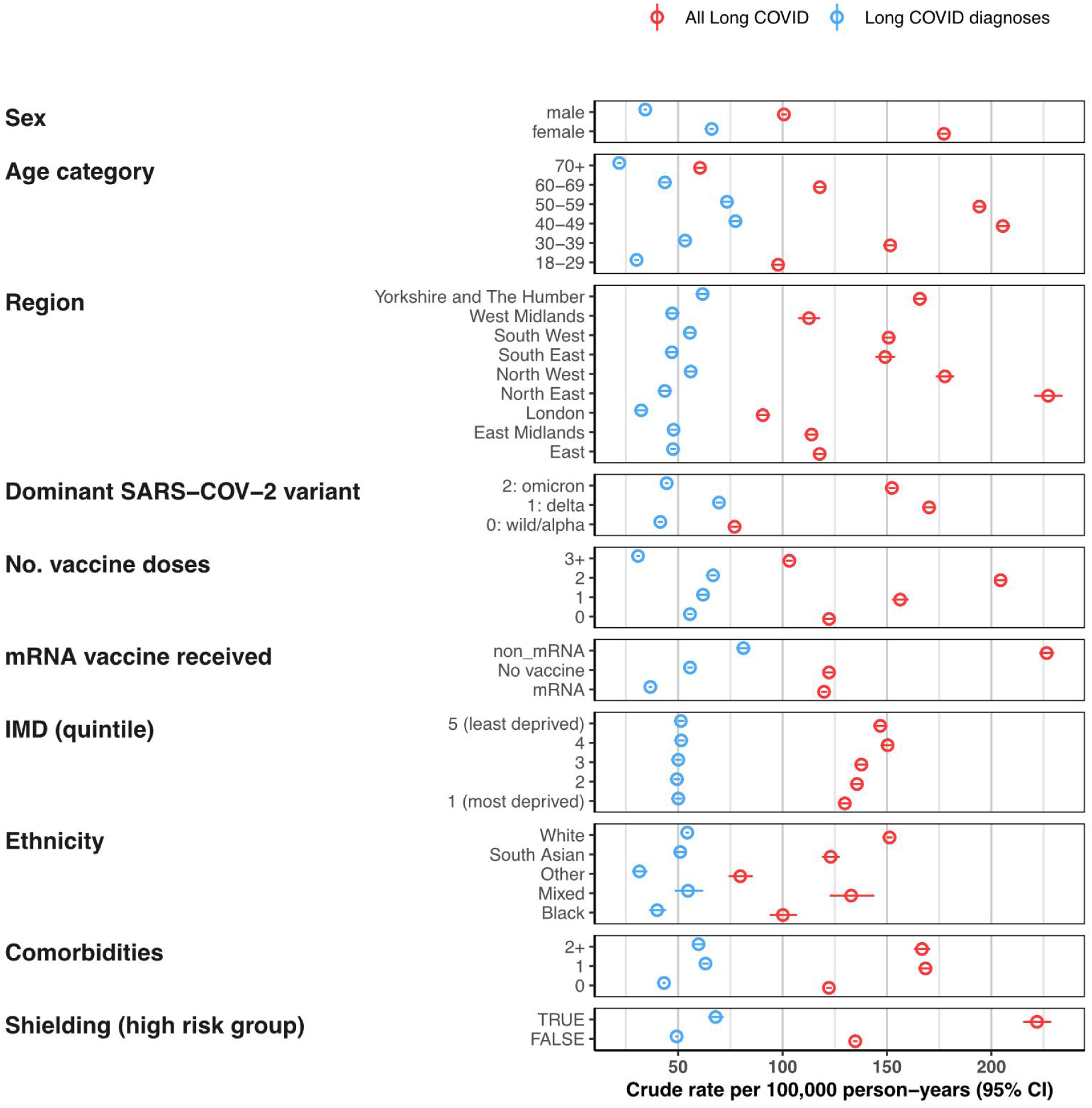
*Rates of EHR recorded long COVID in primary care records per 100,000 person-years. Rate of any long COVID code (red) and long COVID diagnoses only (blue). IMD: index of multiple deprivation*

### Recording long COVID in vaccinated groups

In the primary cohort, followed up until they received a long COVID code, the crude rate of recorded long COVID was lowest for people with 3 or more vaccine doses (103.5 per 100,000 person-years; 95% CI: 101.5-105) (Figure 4**, Table <u>S1</u>**). We estimated the rates in different vaccine dose groups adjusted for age, sex, region and variant, but this is not equivalent to a vaccine effect estimate. The rate of recorded long COVID was 0.85 (95% CI: 0.73-0.99) times lower at least 14 weeks after one vaccine dose, 0.58 (95% CI: 0.5-0.68) times lower after 2 doses, and 0.15 (95% CI: 0.12-0.18) times lower after 3 or more doses, with similar patterns for long COVID diagnosis codes only (Figure 4**, Table <u>S2</u>**). The rate of recording of long COVID is lower in people who received an mRNA-based vaccine for their first dose (RR 0.41; 95% CI: 0.3-0.47) compared to unvaccinated. Those who received adenovirus-based (or other non mRNA formulated vaccines) as a first dose still had lower rates of long COVID in the analysis but with a rate ratio closer to null (0.87; 95% CI: 0.77-0.99). To quality assure the analysis we repeated the models with COVID-19 hospitalisation as an outcome and found associations with age, sex, and vaccination that are consistent with previous research (Figure 4).

**Figure 4:**
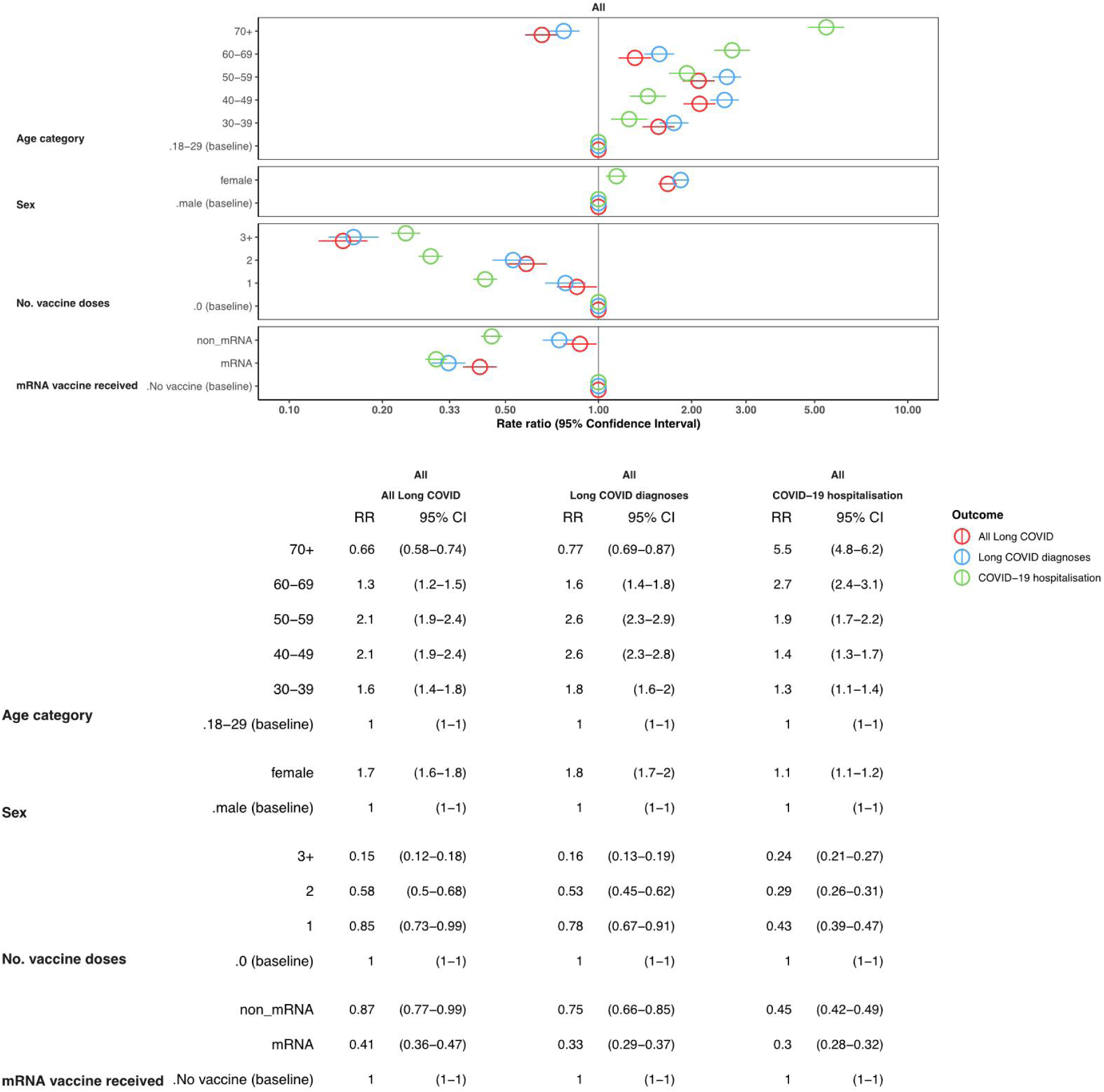
*Minimally-adjusted rate ratios for records of long COVID and COVID-19 hospitalisation. Rate ratios are estimated from negative binomial regression models adjusted for age, sex, 9 NHS regions of England, and the dominant variant circulating*

There is a lag between vaccination and protection from infection, and a further lag until a diagnosis or referral for long COVID can be made. In our main analysis we assumed that this gap is 14 weeks in total. In a sensitivity analysis we expanded this time gap to 18 or 26 weeks. The findings from these results were consistent with our main findings of reduced rates of long COVID with increasing vaccine doses, and a lower rate ratio for those receiving mRNA than non-mRNA vaccines as a first dose (**Figure <u>S7</u>**). We also repeated the analysis stratified by three broad categorisations of the dominant circulating variant of SARS-COV-2 (wild/alpha, delta, omicron). We found that the rate ratios for the effect of vaccination were lowest for long COVID recording during the wild/alpha and delta period and higher during omicron, but were consistently lowest in those with 3+ vaccine doses (**Figure <u>S8</u>**).

In a secondary cohort we continued to follow people after their long COVID record. In this exploratory analysis across we found that the percentage of unvaccinated people was greater in those with a previous long COVID record compared to those without, and this difference was largest in the youngest (18–29) and oldest (70+) age groups (**Figure <u>S9</u>**).

### Differences in long COVID EHR recording routes and relation to SARS-CoV-2 testing

Finally, we investigated the pathways to a long COVID record. We examined the linked SARS-CoV-2 tests and COVID-19 hospitalisation data to calculate the proportion of the 55,465 people with a long COVID record that had previously recorded a positive SARS-CoV-2 test, and been hospitalised with COVID-19. We found that the majority of people with a long COVID record (59%) did not have a recorded positive test result ≥12 weeks before the long COVID record, and a small minority (6.5%) were hospitalised with COVID-19 (Figure 5). There were systematic differences between those with and without a positive test amongst all participants with a long COVID record: those with a previous positive test result were more likely to be female, older, from a more deprived IMD quintile, vaccinated, and to have not been hospitalised with COVID-19 (**Table <u>S3</u>**).

**Figure 5:**
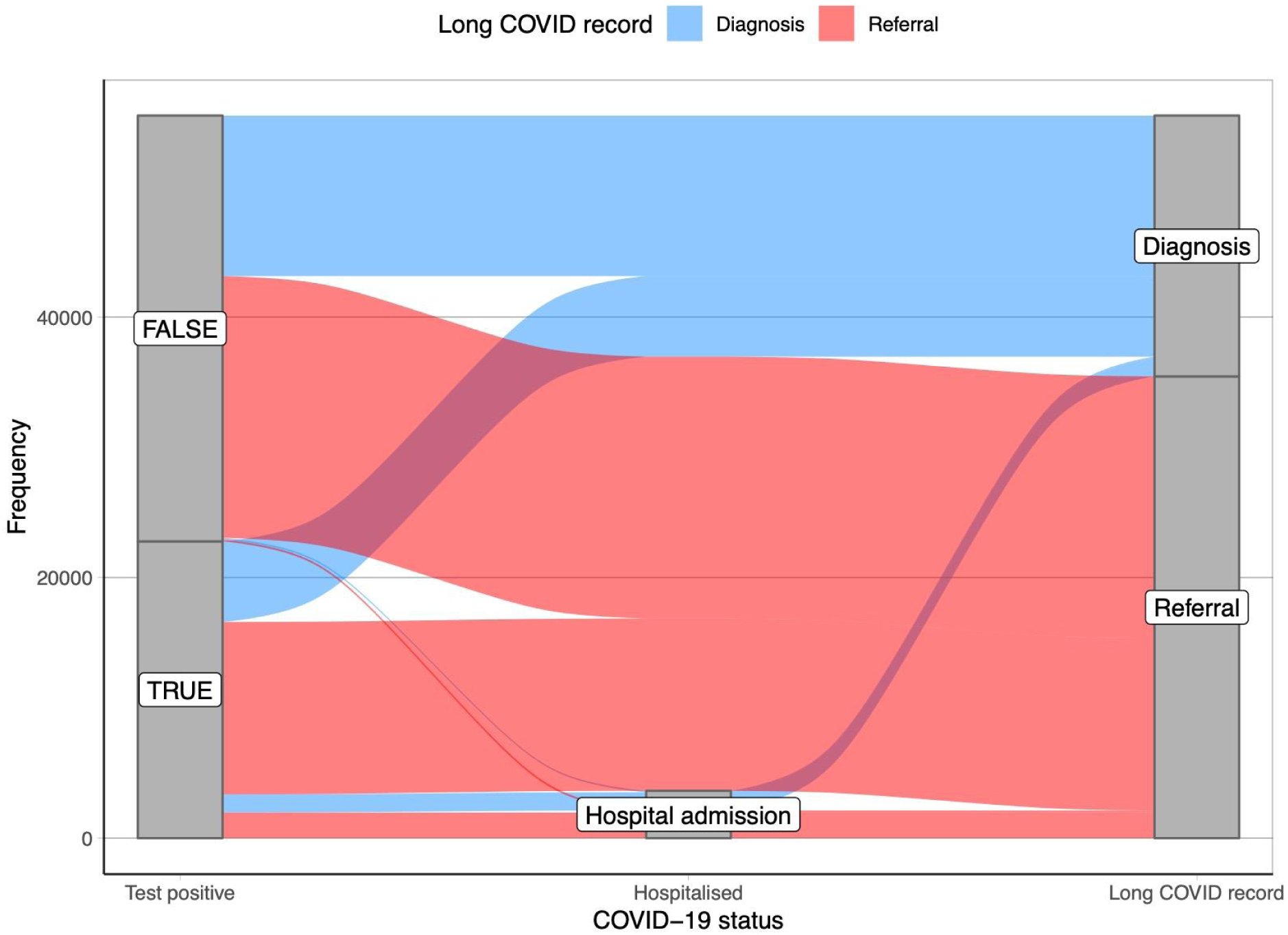
*Sankey diagram of the transition from the presence of a SARS-COV-2 test to a COVID-19 hospitalisation to a long COVID record in primary care for 55,465 participants with a long COVID record*

## Discussion

We analysed the health records of over 19 million adults in England and found very low rates of GP recorded long COVID diagnoses and referrals for long COVID care. We found that referral codes were increasingly common across 2021, but the rate of newly coded patients steadily declined over the year 2022. We found that the choice of long COVID codes used in EHR research to define a long COVID phenotype will have a notable impact on the number of outcomes and temporal dynamics. We do not know if referral or diagnosis codes indicate any difference in severity of symptoms, but there are demographic differences in who received each type of code. Therefore, future studies defining long COVID in these ways should be aware of the different populations represented by each type of code. We also described wide regional variation in long COVID coding and an increase in long COVID referrals in less deprived areas, which may be indicative of greater access to care in these areas (46).

We found that the rate of recording long COVID in primary care was lowest in those with 3 or more vaccine doses, and that those who received an mRNA vaccine initially had lower rates of recorded long COVID than those with an adenovirus-based vaccine. Finally, the majority of people who had a record of long COVID did not have a positive SARS-CoV-2 test at least 12 weeks before the long COVID record, and there were demographic and clinical differences in those with and without a positive test. Therefore, studies that limit to long COVID diagnosis in those with a previous positive test are not selecting patients at random from the recorded long COVID population.

A major strength of our study is the size of the data available. We analysed data on 19 million people and we were able to safely analyse linked primary care, hospitalisation, and SARS-COV-2 test data due to the OpenSAFELY architecture (47). We used a previously defined codelist to identify those with long COVID and to facilitate easy comparisons from research conducted during the pandemic (27). Our analysis has also demonstrated issues with using EHR data for further more complex analyses of long COVID, for example target trial estimation of vaccine efficacy which has been done for other COVID-19 outcomes (38,39).

Our study highlights a large amount of non-differential long COVID misclassification (48). It is likely many people who self-report long COVID in surveys will not have a record from their GP (49). It is also possible that people with a code for long COVID in their GP records do not have the condition, especially those with a referral code as the referral may have concluded that they did not have long COVID. It is also likely that some people will recover from long COVID during this study period which we cannot capture with routine care records. A further limitation is in the timing of long COVID records, which includes all the above limitations plus the systematic differences between people’s propensity to visit their GP with continuing COVID-19 symptoms. This is why we have been explicit throughout the report that we are describing the coding of long COVID, rather than the true incidence of the condition.

Another limitation is possible misclassification of vaccination status and SARS-CoV-2 test status in our cohort. We used a simple method to avoid counting infection events that were not related to the recording of long COVID, by excluding infections <12 weeks, and excluding vaccinations <14 weeks before the end of follow up.This assumes that the recording of long COVID is accurate so that the 12 or 14 week window is accurate. There is an additional possible bias in the recording of positive test results because there may be differences in the rate of false test results over the pandemic (50,51), and because of potential systematic differences in the propensity to record test results between those who do and do not receive a record of long COVID in primary care.

We only had access to practices using SystmOne software, whereas previous work showed that rates of long COVID coding were higher in practices using EMIS software (27). We also assumed that rates were constant over time by using a negative binomial model. This does not allow for changes in the rate that may be directly influenced by the availability of SNOMED codes, changes in clinical guidelines or the availability of long COVID support services.

### Findings in Context

Long COVID codes are rarely recorded in primary care compared to the estimated 2.1 million cases of long COVID self reported in the proactively sampled ONS community infection survey (12). If we assume a crude 10% of SARS-CoV-2 infections result in long COVID, as elsewhere (14), and with approximately 20 million recorded infections in England (42) the number of recorded long COVID cases in primary care is an order of magnitude below the estimated incidence of long COVID in England given the number of SARS-CoV-2 infections. Our findings agree with previous work, that there are serious limitations with simply using EHR records as a measure of long COVID (30,52–54) and alternative approaches may be preferable (28,55). However, our analysis highlights that these other methods may be limited as well, especially if they depend on a recorded positive SARS-CoV-2 test result since we found systematic differences between those with long COVID recorded, with and without a positive test result. The severity of the initial infection may also impact long COVID symptom presentation and potential recording in primary care (56).

The level of long COVID captured in primary care coding is different to other studies, but the temporal trend we found with a decline in incidence over 2022 is consistent with work from the ONS and the USA (12,57). Vaccination and increased natural immunity is a likely contributing factor in this decline, and we found lower rates of long COVID coding with increasing vaccination, which is consistent with other studies (18,58–64). However, there are methodological limitations in previous work as several studies are small (65–67) or in self-selecting populations (68–71) and it is difficult to disentangle the relative contribution of vaccines, variants, and reinfections and how these affect the probability of long-term complications (72–75).

### Policy Implications and Interpretation

As the COVID-19 situation evolves in the UK, surveillance methods are changing and data collection for the ONS COVID-19 infection survey was paused in March 2023. This limits the possible data sources for monitoring and understanding long COVID. We have shown that long COVID clinical coding is limited in comparison to nationally representative random sampling such as the ONS-CIS, but EHRs have the potential to be an important resource for long COVID research. However, until we can better understand the reasons for the under-reporting of cases in primary care, this potential will not be realised.

One attractive solution in EHRs is to develop an alternative algorithm of detection method from rich data for identifying people with long COVID through identification of symptoms associated with long COVID (28,29,76). These methods often include a positive SARS-CoV-2 test result as a prerequisite, however our work shows that this would fail to detect 59% of the recorded long COVID coded individuals in our cohort. A combination of detection methods is therefore a necessity in future EHR long COVID research.

### Future Research

Data from multiple sources is needed to validate the definitions of long COVID between studies and establish a consistent definition so that research findings are generalisable outside of a specific study with a specific outcome definition. Validation of outcome measures is needed to better capture cases. Future research should combine routinely collected data with more granular detailed survey responses (12,75,77) to better understand the differences between these data sources and triangulate evidence.

Despite the difficulties in researching vaccine effectiveness on long COVID, it is an important question to understand. It is unclear what role vaccination had in the protection against long COVID, beyond reduced risk of any infection. Further analysis could expand on research of heterogeneous vaccine mixing and different vaccine schedules and the impact these had on infections (78–81), and whether these possible benefits confer to reduced long term COVID-19 incidence or symptom burden.

### Summary

Many people in the UK suffer with long COVID following the COVID-19 pandemic but relatively few cases are recorded in primary care. There are many uncertainties about long COVID (82,83) and EHRs have the potential to shed light on these. However, more work is needed on the definition and identification of a “case of long COVID” before this can be realised.

#### Abbreviations

EHR: Electronic Health Records
SARS-COV-2: Severe acute respiratory syndrome coronavirus 2

## Data Availability

Access to the underlying identifiable and potentially re-identifiable pseudonymised electronic health record data is tightly governed by various legislative and regulatory frameworks, and restricted by best practice. The data in the NHS England OpenSAFELY COVID-19 service is drawn from General Practice data across England where TPP is the data processor.
TPP developers initiate an automated process to create pseudonymised records in the core OpenSAFELY database, which are copies of key structured data tables in the identifiable records. These pseudonymised records are linked onto key external data resources that have also been pseudonymised via SHA-512 one-way hashing of NHS numbers using a shared salt. University of Oxford, Bennett Institute for Applied Data Science developers and PIs, who hold contracts with NHS England, have access to the OpenSAFELY pseudonymised data tables to develop the OpenSAFELY tools.
These tools in turn enable researchers with OpenSAFELY data access agreements to write and execute code for data management and data analysis without direct access to the underlying raw pseudonymised patient data, and to review the outputs of this code. All code for the full data management pipeline - from raw data to completed results for this analysis - and for the OpenSAFELY platform as a whole is available for review at github.com/OpenSAFELY.

## Administrative

## Acknowledgements

We are very grateful for all the support received from the TPP Technical Operations team throughout this work, and for generous assistance from the information governance and database teams at NHS England and the NHS England Transformation Directorate.

## Conflicts of Interest

All authors have completed the ICMJE uniform disclosure form at www.icmje.org/coi_disclosure.pdf and declare the following:…

## Funding

This research was supported by the National Institute for Health and Care Research (NIHR) (OpenPROMPT: COV-LT2-0073)). In addition, this research used data assets made available as part of the Data and Connectivity National Core Study, led by Health Data Research UK in partnership with the Office for National Statistics and funded by UK Research and Innovation (grant ref MC_PC_20058). In addition, the OpenSAFELY Platform is supported by grants from the Wellcome Trust (222097/Z/20/Z); MRC (MR/V015737/1, MC_PC-20059, MR/W016729/1); NIHR (NIHR135559, COV-LT2-0073), and Health Data Research UK (HDRUK2021.000, 2021.0157).

The views expressed are those of the authors and not necessarily those of the NIHR, NHS England, UK Health Security Agency (UKHSA) or the Department of Health and Social Care.

Funders had no role in the study design, collection, analysis, and interpretation of data; in the writing of the report; and in the decision to submit the article for publication.

## Information governance and ethical approval

NHS England is the data controller of the NHS England OpenSAFELY COVID-19 Service. TPP is the data processor; all study authors using OpenSAFELY have the approval of NHS England. This implementation of OpenSAFELY is hosted within the TPP environment which is accredited to the ISO 27001 information security standard and is NHS IG Toolkit compliant. Patient data has been pseudonymised for analysis and linkage using industry standard cryptographic hashing techniques; all pseudonymised datasets transmitted for linkage onto OpenSAFELY are encrypted; access to the NHS England OpenSAFELY COVID-19 service is via a virtual private network (VPN) connection; the researchers hold contracts with NHS England and only access the platform to initiate database queries and statistical models; all database activity is logged; only aggregate statistical outputs leave the platform environment following best practice for anonymisation of results such as statistical disclosure control for low cell counts.

The service adheres to the obligations of the UK General Data Protection Regulation (UK GDPR) and the Data Protection Act 2018. The service previously operated under notices initially issued in February 2020 by the the Secretary of State under Regulation 3(4) of the Health Service (Control of Patient Information) Regulations 2002 (COPI Regulations), which required organisations to process confidential patient information for COVID-19 purposes; this set aside the requirement for patient consent. As of 1 July 2023, the Secretary of State has requested that NHS England continue to operate the Service under the COVID-19 Directions 2020. In some cases of data sharing, the common law duty of confidence is met using, for example, patient consent or support from the Health Research Authority Confidentiality Advisory Group.

Taken together, these provide the legal bases to link patient datasets using the service. GP practices, which provide access to the primary care data, are required to share relevant health information to support the public health response to the pandemic, and have been informed of how the service operates.

This research is part of the OpenPROMPT study “Quality-of-life in patients with long COVID: harnessing the scale of big data to quantify the health and economic costs” which has ethical approval from HRA and Health and Care Research Wales (HCRW) (IRAS project ID 304354). The Study Coordination Centre has obtained approval from the LSHTM Research Ethics Committee (ref 28030), as well as a favourable opinion from the South Central—Berkshire B Research Ethics Committee (ref 22/SC/0198).

## Data access and verification

Access to the underlying identifiable and potentially re-identifiable pseudonymised electronic health record data is tightly governed by various legislative and regulatory frameworks, and restricted by best practice. The data in the NHS England OpenSAFELY COVID-19 service is drawn from General Practice data across England where TPP is the data processor.

TPP developers initiate an automated process to create pseudonymised records in the core OpenSAFELY database, which are copies of key structured data tables in the identifiable records. These pseudonymised records are linked onto key external data resources that have also been pseudonymised via SHA-512 one-way hashing of NHS numbers using a shared salt. University of Oxford, Bennett Institute for Applied Data Science developers and PIs, who hold contracts with NHS England, have access to the OpenSAFELY pseudonymised data tables to develop the OpenSAFELY tools.

These tools in turn enable researchers with OpenSAFELY data access agreements to write and execute code for data management and data analysis without direct access to the underlying raw pseudonymised patient data, and to review the outputs of this code. All code for the full data management pipeline — from raw data to completed results for this analysis — and for the OpenSAFELY platform as a whole is available for review at github.com/OpenSAFELY.

The data management and analysis code for this paper was led by ADH and contributed to by BB-C.

## Notes

### Competing Interest Statement

BG is a Non-Executive Director at NHS Digital; he also receives personal income from speaking and writing for lay audiences on the misuse of science. All other authors declare no competing interests.

### Author Declarations

This research is part of the OpenPROMPT study 'Quality-of-life in patients with long COVID: harnessing the scale of big data to quantify the health and economic costs' Health Research Authority (HRA) and Health and Care Research Wales (HCRW) gave ethical approval for this work (IRAS project ID 304354). The Research Ethics Committee of the London School of Hygiene & Tropical Medicine gave ethical approval for this work (ref 28030) Research Ethics Committee of South Central-Berkshire B gave favourable opinion (ref 22/SC/0198).

